# Biologic Therapies for the Treatment of Post Ileal Pouch Anal Anastomosis Surgery Chronic Inflammatory Disorders: Systematic Review and Meta-analysis

**DOI:** 10.1101/2021.05.22.21257585

**Authors:** Mohammad Shehab, Fatema Alrashed, Aline Charabaty, Talat Bessissow

## Abstract

**Background:** Chronic inflammatory disorders after ileal pouch-anal anastomosis (IPAA) surgery are common. These include chronic pouchitis, Crohn’s disease of the pouch, prepouch ileitis, and rectal cuff inflammation. The aim of this study was to evaluate the efficacy of biologic therapies in treating these disorders.

**Method:** Systematic review of all published studies from inception to April 1st, 2021 was performed to investigate the efficacy of biologic therapies for post IPAA chronic inflammatory disorders. The primary outcome was the efficacy of biologic therapies in achieving complete response or remission. A subgroup analysis was performed for each disorder separately.

**Results:** A total of 24 studies were identified including 682 patients. Using a random effect model, the overall pooled efficacy of biologic therapies in achieving complete response or remission in patients with post IPAA surgery chronic inflammatory disorders was 60% (95% Confidence Interval (CI), 56 - 64). Specifically, the pooled efficacy of ustekinumab was 75% (95% CI, 64 - 85, p = 0.007), whereas the efficacy of vedolizumab was 60% (95% CI, 52 - 68, p = 0.172). In addition, the efficacy infliximab and adalimumab were 59% (95% CI, 53 - 64, p <0.01) and 51% (95% CI, 42 - 60, p = 0.452) respectively.

**Conclusion:** Ustekinumab, infliximab, vedolizumab and adalimumab are effective in achieving complete response or remission in post IPAA surgery chronic inflammatory disorders. There is also a clear trend toward higher efficacy in patients with Crohn’s disease of the pouch compared to chronic pouchitis. More studies are needed to determine the efficacy of biologics in cuffitis.

**Summary:** Chronic inflammatory disorders after ileal pouch-anal anastomosis surgery are common. These include chronic pouchitis, Crohn’s disease of the pouch, prepouch ileitis, and rectal cuff inflammation. This study aimed to evaluate the efficacy of biologic therapies in achieving complete response or remission in these disorders.

## Introduction

Proctocolectomy remains the therapy of choice for patients with ulcerative colitis (UC) who are refractory to medical therapy, develop unresectable colonic dysplasia or colorectal malignancy.^1-3^ The lifetime risk of colectomy in patients with UC remains between 10-30% despite the era of biologic therapy.^4^ Proctocolectomy followed by ileal pouch anal anastomosis (IPAA) is one of the preferred surgical approaches^5^. However, chronic inflammatory complications following (IPAA) surgery are not uncommon.^6-7^

Chronic pouchitis (CP), Crohn’s disease of the pouch (CD), prepouch ileitis (PI), and rectal cuff inflammation (cuffitis) are the most common chronic inflammatory disorders post IPAA. In addition, the pathophysiology of each of these conditions is different, hence the treatment is also different. Apart from conventional treatment with antibiotics, 5-ASAs or steroids, biologic therapies have been used to treat these conditions that are refractory to initial therapy. However, the efficacy of each biologic for each of these conditions remains unclear. We performed a systematic review and meta-analysis to better understand the magnitude of the effect of each biologic therapy to tailor our practice accordingly.

## Methods

This systematic review and meta-analysis were conducted using the methods described in the Cochrane Handbook of Systematic Reviews and reported according to the PRISMA (Preferred Reporting Items for Systematic Reviews and Meta-Analyses) statement.^9^ MOOSE guidelines were also followed (see attached supplementary checklists).^10^

### Eligibility Criteria

Randomized, placebo-, or active comparator-controlled trials, cohort studies, observational studies and case series reporting biologic therapies use post IPAA surgery were included. Adult patients (age ≥18 years) with post IPAA chronic inflammatory disorders who were refractory to 4 weeks of conventional therapy were included. Our study focus is biological pharmacological treatments alone or in combination with other agents. We excluded trials studying only pediatric patients (age, <18 years), case reports, editorials, and correspondences, studies that did not evaluate a biological agent and those where data could not be extracted.

### Definitions

For the purpose of this study, chronic pouchitis, Crohn’s disease of the pouch, prepouch ileitis, and rectal cuff inflammation were considered to be post IPAA chronic inflammatory disorders. Chronic pouchitis (CP) was defined as inflammation of the pouch refractory to 4 weeks of first line therapy (antibiotics, 5-ASAs and steroids) whereas Crohn’s disease of the pouch (CD) was defined as the presence of one of the following: non-anastomotic fistula involving the perineum or small bowel, non-anastomotic stricture of pouch, skip ulcerations of the pouch with or without extension to the afferent limb, or histological features suggestive of CD. Prepouch ileitis (PI) was defined as chronic inflammation of the afferent limb of the pouch.

### Search Strategy, Data Extraction and Outcomes

Literature searches were conducted by two authors (MS and FA) using MEDLINE, Embase, Scopus, and Cochrane Central Register of Controlled Trials databases were searched from inception to April 1st, 2021 using predefined strategies (Supplementary Appendix 1). Our search strategies were designed with the help of a librarian. The search was restricted to English language publications involving humans. English conference proceedings were searched [World Congress of Gastroenterology, American College of Gastroenterology, Canadian Digestive Disease Week, Digestive Disease Week, European Crohn’s and Colitis Organization congress, and United European Gastroenterology Week]. Furthermore, clinical trials databases [www.clinicaltrials.gov and International Randomized Standard Clinical Trial (IRSCT) Register] were searched. Google scholar was also searched for unindexed trials. The search terms used are outlined in the supplemental material.

Data extraction and quality control were done independently by two reviewers (MS and FA). Discrepancies were resolved by a third reviewer (TB). The same two authors extracted information from the studies. Extracted information included baseline characteristics, study design, risk of bias, year and country of publication, intervention, outcomes and mean follow up duration using standardized excel spreadsheet.

The primary outcome measure was the efficacy of biologics in inducing complete response or remission, as defined by each study, after failure of 4 weeks of first line therapy.

### Statistical Analysis

Meta-analysis methods were used to pool the percentage of patients who achieved complete response or remission from the various studies. Statistical analysis was conducted using Stata software (version 13; StataCorp, College Station, TX, USA). Prevalence and 95% confidence interval were estimated using random-effects models assuming between and within study variability.

### Heterogeneity analysis

I^2^ statistic, which ranges from 0% to 100%, was used to quantify the relative amount of observed heterogeneity. An I^2^ value less than 30% indicates low heterogeneity, whereas a range of 30 - 75% indicates moderate heterogeneity and high heterogeneity was defined as I^2^ > 75%. Sources of heterogeneity were explored by performing multiple sensitivity analyses.

### Subgroup Analysis

To explore the effect of different biologics on specific post IPAA chronic inflammatory disorders, a subgroup analysis was performed. It included patients with: [i] chronic pouchitis [ii] Crohn’s diease of the pouch and [iii] prepouch ileitis. Cuffitis was not analyzed as no study was found that matched our inclusion criteria.

### Sensitivity Analysis

To identify potential sources of heterogeneity, 3 different sensitivity analyses were performed; [i] location where study was performed [within North America compared to outside countries [ii] Quality of studies: moderate to high quality studies were only included, defines as modified Newcastle-Ottawa Scale^11^ (mNOS) of 4 and above and [iii] study published after May 2014, as vedolizumab and ustekinumab were approved by U.S. Food and Drug Administration (FDA) after that time period. It is possible that patients on these medications may have received anti TNF therapy, i.e are anti TNF experience.

### Risk of Bias and Study Quality

To assess risk of bias and quality of the included studies, two authors (MS and FA) independently used the Cochrane risk of bias tool for randomized controlled trial and ROBINS-I for assessing risk of bias in non-randomized studies of interventions.^12^ By using these assessment tools, studies were classified as being of unclear or low or high risk of bias. Seven domains: random sequence generation, allocation concealment, blinding of participants and personnel, blinding of outcome assessment, incomplete outcome data, selective outcome reporting, and other potential sources of bias are included in this tool.

The quality of all included studies were assessed using the modified Newcastle-Ottawa Scale (mNOS). Three domains were assessed by using mNOS: selection, compatibility and outcome. Study quality was defined as low (score of 0-3), moderate (score of 4-6) and High (score of 7 and 8).

## Results

### Search Results

Our search identified 611 studies (figure 1). After exclusion of duplicates and selection based on our prespecified inclusion and exclusion criteria, twenty-four studies^13-36^ were included in the main analysis. All studies were observational except for one, which was a randomized control trial. Twelve studies were published in North America and the rest were published in different countries in Europe.

**Figure 1.**
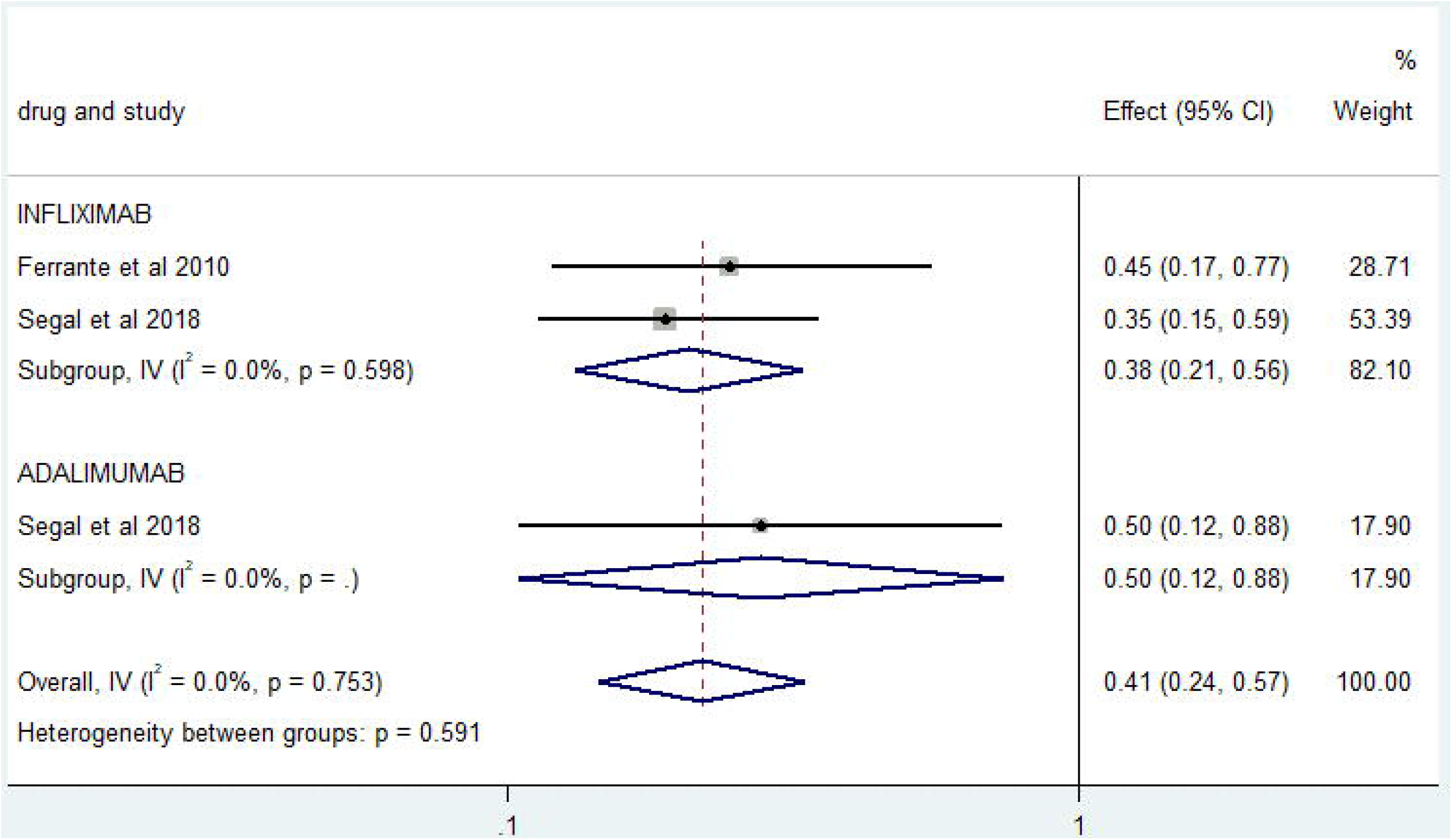
PRISMA flow chart outlining the search process for selecting the studies included in this systematic review with meta-analysis

### Patient Characteristics

The selected studies included a total of 682 patients diagnosed with one of the post IPAA chronic inflammatory disorders (see supplemental table 2). Mean age was 35 (±7), and 366 (53.7%) participants were males. The mean disease duration was 9.9 years, whereas the mean follow up duration was 16.4 months. 269 patients were treated with infliximab, 143 patients with vedolizumab, 164 patients with adalimumab and 66 patients with ustekinumab. The rest received different non-biologic treatments (controls) or were lost to follow up.

### Main Outcomes

Using a random effect model, the efficacy of ustekinumab in achieving complete response or remission in patients with chronic inflammatory disorders post IPAA surgery was 75% (95% CI, 64 - 85, p = 0.007), whereas the efficacy of vedolizumab was 60% (95% CI, 51 - 68, p = 0.172). The Efficacies of Infliximab and adalimumab were 59% (95% CI, 53 - 64, p <0.01) and 51% (95% CI, 42 - 60, p = 0.452) respectively. This analysis showed a moderate heterogeneity between the different studies, with I^2^ value of 62.7% (see figure 2).

**Figure 2.**
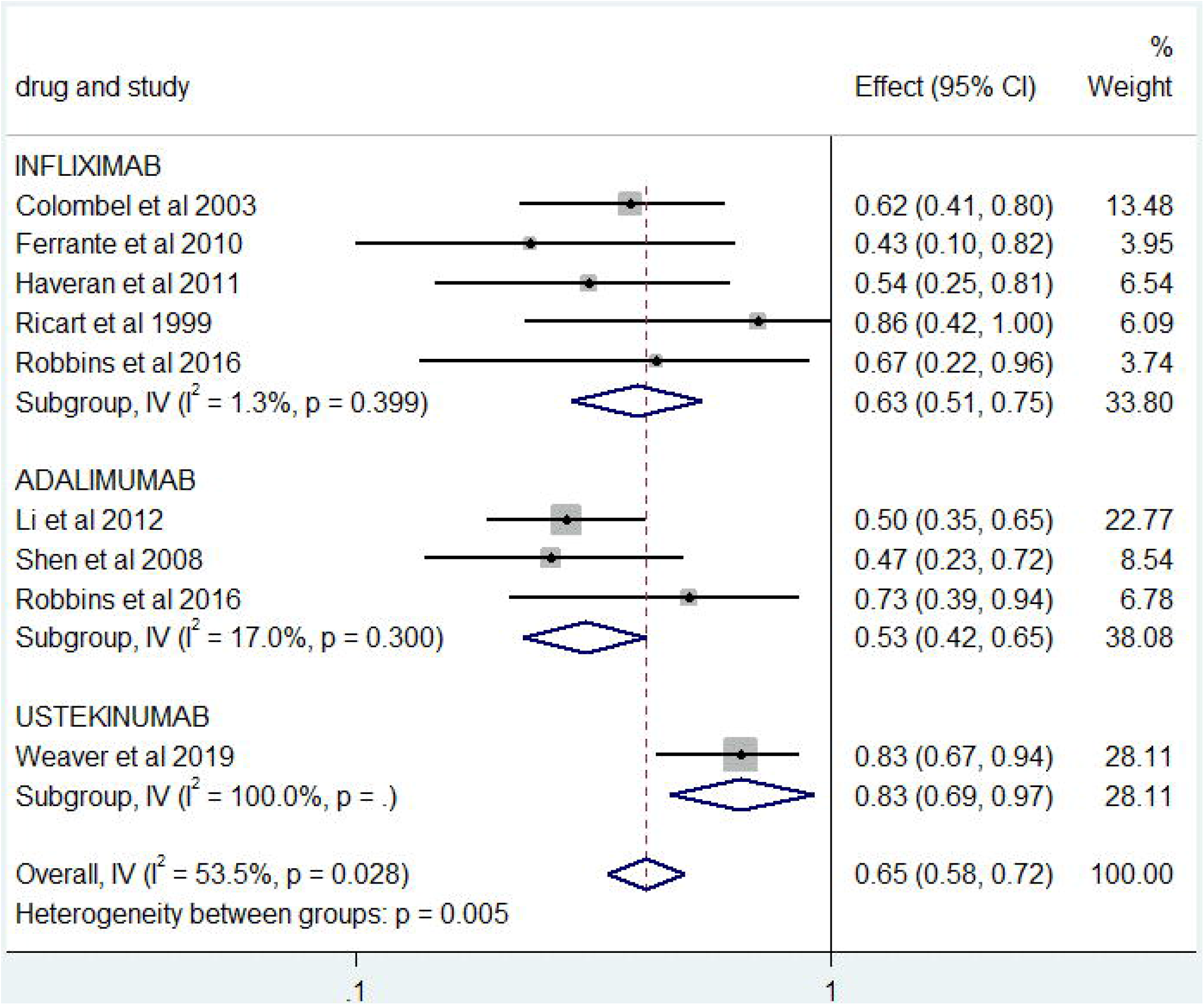
Forest plot showing the overall efficacy of biologic therapies in achieving complete response or remission in patients with inflammatory disorder post IPAA surgery

### Subgroup Analysis

Our subgroup analysis showed that the efficacy of biologics in achieving complete response or remission in patients with chronic pouchitis was 59% (95% CI, 54 - 63, p <0.01) (figure 3). In patients with Crohn’s disease of the pouch, biologic therapy efficacy was higher at 65% (95% CI, 58 - 72, p <0.01) (figure 4). Whereas in patients with prepouch ileitis the efficacy was 41% (95% CI, 24 - 57, p = 0.591) (figure 5).

**Figure 3.**
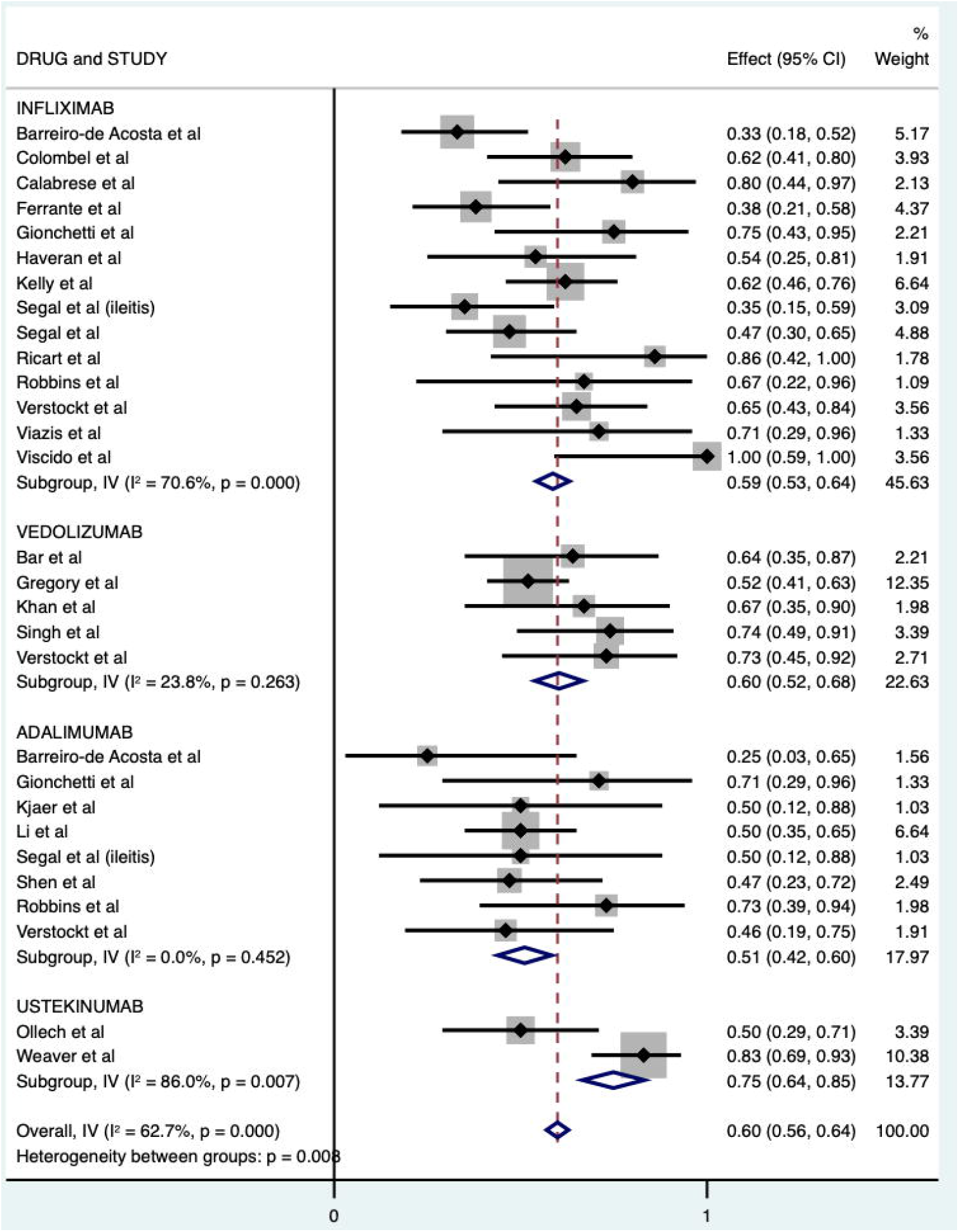
Forest plot showing the efficacy of biologic therapies in achieving complete response or remission in patients with chronic pouchitis

**Figure 4.**
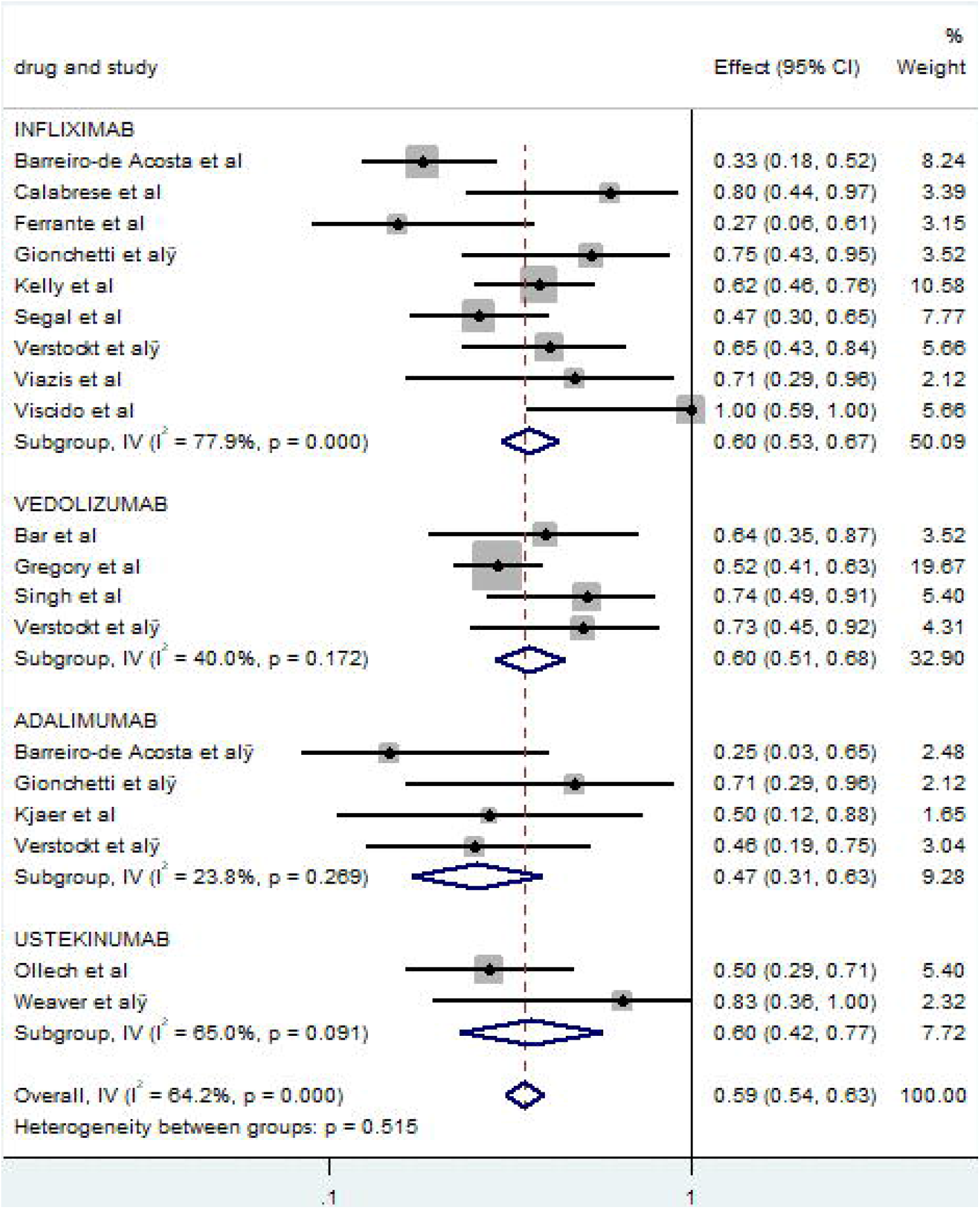
Forest plot showing the efficacy of biologic therapies in achieving complete response or remission in patients with Crohn’s disease of the pouch

**Figure 5.**
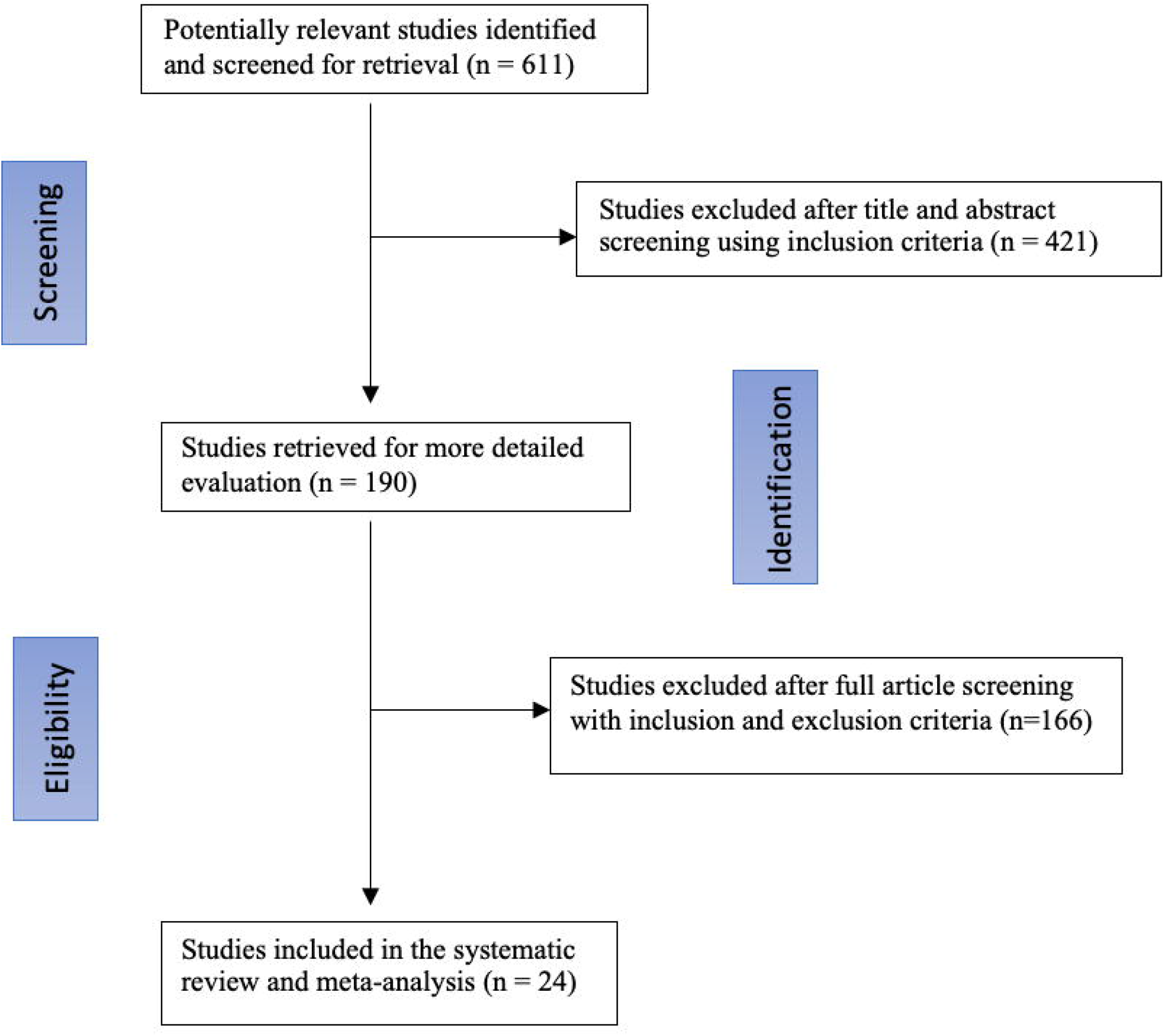
Forest plot showing the efficacy of biologic therapies in achieving complete response or remission in patients with prepouch ileitis

When looking at each drug individually, the pooled efficacy of infliximab in chronic pouchitis was 60% (95% CI, 53 - 67, p <0.01), whereas the efficacies of vedolizumab, adalimumab and ustekinumab were 60% (95% CI, 51 - 68), 47% (95% CI, 31 - 63), and 60% (95% CI, 54 - 63) respectively (figure 3). In patients with CD of the pouch, the efficacy of infliximab, adalimumab and ustekinumab in achieving complete response or remission was 63% (95% CI, 51 - 75), 53% (95% CI, 42 - 65) and 83% (95% CI, 69 - 97) (figure 4). With regards to prepouch ileitis, the included studies investigated infliximab and adalimumab. The efficacy of infliximab in achieving complete response or remission in patients with prepouch ileitis was 38% (95 % CI, 21 - 56), whereas the efficacy of adalimumab was 50% (95% CI, 12-88) (figure 5).

### Sensitivity Analysis

After restricting the analysis to studies performed in North America, study period, or high quality studies the pooled efficacy of individual biologic therapies did not differ substantially as described below:

#### 1) North America only

pooled efficacy of biologic therapies in achieving complete response or remission in patients with chronic inflammatory disorders of pouch was 63% (95% CI, 54 - 71). Specifically, the pooled efficacy of ustekinumab in achieving complete response or remission was 68% (95% CI, 35 - 1.00), whereas the efficacy of infliximab was 64% (95% CI, 54 - 74). In addition, the efficacy of vedolizumab and adalimumab were 61% (95% CI, 40 - 82) and 54% (95% CI, 41 - 67) respectively (see supplemental figure 1).

#### 2) Moderate or high quality studies only

after removing the low quality studies (mNOS of 3 or less), pooled efficacy of biologic therapies in achieving complete response or remission in patients with chronic inflammatory disorders of pouch was 54% (95% CI, 64 - 63). Specifically, the pooled efficacy of ustekinumab in achieving complete response or remission was 68% (95% CI, 35 - 1.00), whereas the efficacy of vedolizumab was 54% (95% CI, 44 - 64). In addition, the efficacy infliximab and adalimumab were 52% (95% CI, 40 - 64 and 48% (95% CI, 34 - 62) respectively (see supplemental figure 2).

#### 3) Studies published after May 2014 only

Sensitivity analysis performed based on study period showed that pooled efficacy of biologic therapies in achieving complete response or remission in patients with chronic inflammatory disorders of pouch was 60% (95% CI, 52 - 67). Specifically, the pooled efficacy of ustekinumab in achieving complete response or remission was 68% (95% CI, 35 - 1.00), whereas the efficacy of vedolizumab was 63% (95% CI, 51 - 75). In addition, the efficacies infliximab and adalimumab were 54% (95% CI, 45 - 64) and 56% (95% CI, 41 - 72) respectively (see supplemental figure 3).

### Study quality and risk of bias

Median mNOS score was 4, with scores ranging from 2 to 8 (see supplemental table 3). In terms of risk of bias, most studied were judged to have low risk of bias using the Cochrane risk of bias tool for randomized controlled trial and ROBINS-I, while six studies had moderate risk of bias (see supplemental table 4).

### Publication bias

Supplemental figure 4 shows a funnel plot of publication bias. Based visual examination of the plot, symmetrical distribution of the studies on the funnel plot suggests no publication bias.

## Discussion

Chronic pouchitis (CP) is the most common complication after IPAA, affecting approximately 40% of patients in the first years after surgery.^37^ This was reflected in our study as well where 47% of total patients included had CP. A recent study evaluated the burden of inflammatory conditions of the pouch in a tertiary hospital and found that up to 30% of patients developed chronic pouchitis that required reinitiation of biologic therapy.^38^ In addition, patients with Crohn’s disease of the pouch (CD) were found to require early biologic therapy soon after IPAA to prevent further complications.^39^ This emphasizes the importance of understanding the effectiveness of the different biologic therapies in inducing complete response or remission in these patients.

In this meta-analysis the overall efficacy of biologics in achieving complete response or remission in patients with inflammatory disorders of the pouch was 60% (95% CI, 56 - 64) and the efficacy of infliximab was 59% (95% CI, 53 - 64). These results are similar to the findings of the meta-analysis by Haguet et al. In our study, adalimumab efficacy was 51% (95% CI, 42 - 60), which was higher than Haguet et al pooled efficacy of adalimumab at 30%. This could be due to several reasons: our meta-analysis included a higher number of studies, and included studies of biologics for prepouch ileitis, whereas Haguet et al meat-analysis was limited to studies with diagnosis of chronic pouchitis and Crohn’s disease of the pouch. Another meta-analysis explored the effectiveness of multiple therapies in achieving remission in patients with chronic pouchitis, and found an overall remission rate with biologic therapy of 53%.^41^ This study also suggested that antibiotics have higher efficacy in the treatment of chronic pouchitis. While antibiotics are generally considered first line therapy, the treatment of pouchitis is often a challenge and requires multiple therapies in order to achieve remission.

In addition to the effect of biologics on chronic pouchitis, our study explored the efficacy of biologics in other inflammatory disorders of the pouch including prepouch ileitis (PI) and Crohn’s disease of the pouch. One study found that prepouch ileitis was independently associated with an increased risk of Crohn’s disease like complications, need for surgical/endoscopic intervention, and immunosuppressive therapy.^42^ These findings underline the strong association of PI with disease complications and emphasize the need to treat early with the most effective therapy. Patients with PI accounted for 5.4% of the total patients included in our meta-analysis, a number similar to what was seen in two other studies that reported a 6% frequency of this complication among patients with IPAA ^43-44^ Our subgroup analysis found that the pooled efficacy of biologic therapy in patients with PI is 41% (95% CI, 24 - 57). Prepouch ileitis (PI) in patients with IPAA is an uncommon occurrence and finding a large group of such patients to study is challenging, especially at a single center.

To the best of our knowledge this is the only systematic review and meta-analysis that explored biologic therapies efficacy in all inflammatory disorders post IPAA surgery. In addition, this study is the first to investigate multiple biologic therapies other than anti-TNF agents. This study however has some limitations. `All studies, except one, were observational studies, with 17 out of 24 studies performed retrospectively. This can lead to high risk of confounding and selection bias as well as high heterogeneity among the studies. To limit this, we performed multiple sensitivity analyses. In addition, each study definition of response or remission was different, which ranged from symptoms resolution to change in the Pouchitis Disease Activity Index (PDAI). We addressed this limitation in our study by including complete response or remission as defined by each study, rather than partial response or remission. Furthermore, the lack of data on patients with cuffitis made it difficult to draw any conclusion regarding the efficacy of biologic therapies in this population of patients.

### Conclusion

Ustekinumab, infliximab, vedolizumab and adalimumab are effective in achieving complete response or remission post IPAA chronic inflammatory disorders. There is also a clear trend toward higher efficacy in patients with Crohn’s disease of the pouch compared to chronic pouchitis. Randomized control trials are needed to determine precise efficacy of biologics in patients with chronic inflammatory disorders post IPAA surgery. In addition, studies needed to determine the efficacy of biologics in cuffitis.

## Supporting information

PRISMA CHECKLIST

MOOSE CHECKLIST

## Data Availability

All research data available with the submitted documents.

## Acknowledgment

none.

