## supplementary documents for "Biologic Therapies for the Treatment of Post Ileal Pouch Anal Anastomosis Surgery Chronic Inflammatory Disorders: Systematic Review and Meta-analysis"

### Search Strategy

| Supplemental Table 1 \| Search strategy |
|---|
| **Medline search strategy for Refractory Pouchitis and prepouch ileitis** |
| ("Pouchitis"[Mesh] or pouchitis[tw] or Pouch Ileitis[tw] or Prepouch Ileitis[tw] or "colonic pouches"[Mesh] or colonic pouches[tw] or Pelvic Pouches[tw] or Pelvic Pouches [tw] or W Pouch[tw] or Ileal Pouche[tw] or J Pouch[tw] or s-pouch [tw] or IPAA [tw])<br>AND<br>("Biologics"[Mesh] or Biologic*[tw] or Biopharmaceutical*[tw] or Biological Drug*[tw] or Biological*[tw] or Biologic Medicine*[tw] or Biologic Pharmaceutical*[tw] or Biologic Drug*[tw] or Biological Medicine[tw] or "Monoclonal Antibodies"[Mesh] or Monoclonal Antibody*[tw] or anti-TNF[tw] or "Tumor Necrosis Factor alpha"[Mesh] or Tumor Necrosis Factor alpha[tw])<br>AND<br>("Therapy"[Mesh] or therapy[tw] or treatment[tw] or "remission induction"[Mesh] or remission induction[tw]) |
| **Medline search strategy for Crohn's disease of the pouch** |
| ("Colonic Pouche"[Mesh] or Colonic Pouche[tw] or Ileoanal Pouche*[tw] or Ileoanal Reservoir [tw] or Pelvic Pouche*[tw] or Pelvic Pouche*[tw] or W Pouch[tw] or Ileal Pouche[tw] or J Pouch[tw] or s-pouch[tw] or IPAA[tw])<br>AND<br>("Biologics"[Mesh] or Biologic*[tw] or Biopharmaceutical*[tw] or Biological Drug*[tw] or Biological*[tw] or Biologic Medicine*[tw] or Biologic Pharmaceutical*[tw] or Biologic Drug*[tw] or Biological Medicine[tw] or "Monoclonal Antibodies"[Mesh] or Monoclonal Antibody*[tw] or anti-TNF[tw] or "Tumor Necrosis Factor alpha"[Mesh] or Tumor Necrosis Factor alpha[tw])<br><br>AND |

("Therapy"[Mesh] or therapy[tw] or treatment[tw] or "remission induction"[Mesh] or remission induction[tw])

**Medline search strategy for Cuffitis**

(Rectal cuff inflammation*[tw] or cuffitis*[tw])

AND

("Biologics"[Mesh] or Biologic*[tw] or Biopharmaceutical*[tw] or Biological Drug*[tw] or Biological*[tw] or Biologic Medicine*[tw] or Biologic Pharmaceutical*[tw] or Biologic Drug*[tw] or Biological Medicine[tw] or "Monoclonal Antibodies"[Mesh] or Monoclonal Antibody*[tw] or anti-TNF[tw] or "Tumor Necrosis Factor alpha"[Mesh] or Tumor Necrosis Factor alpha[tw])

AND

("Therapy"[Mesh] or therapy[tw] or treatment[tw] or "remission induction"[Mesh] or remission induction[tw])

Supplemental Table. 2

| Study | Number of patients | Gender (male) n | Mean age | Disease duration before IPAA (years) | Mean follow up after IPAA (months) | Outcomes | Study type |
|---|---|---|---|---|---|---|---|
| Bar et al | 20 | 12 | 22.5 | 3.5 | Not reported | The effectiveness was measured using the Oresland Score (OS) at week 2, 6, 10 and 14 and the pouch disease activity index (PDAI) at week 0 and 14. | Retrospective |
| Calabrese et al | 10 | 6 | 39.2 | 6.3 | 7.3 | Macroscopic remission was defined as the disappearance of all large lesions and the persistence of £3 small lesions, at WCE and pouch endoscopy. | Prospective |
| Colombel et al | 26 | 7 | 32 | 4 | 21.5 | The main clinical outcomes for long term response were pouch excision and presence of diverting ileostomy. | retrospective |

| | | | | | | | |
|---|---|---|---|---|---|---|---|
| Haveran et al | 22 | 14 | 27.8 | 8.9 | 97 | Success in treatment was defined as complete resolution or significant improvement of symptoms specific to the indication for therapy | Retrospective |
| Li et al | 48 | 29 | 25.3 | 4 | 25 | Complete clinical response was defined as the resolution of symptoms as well as the modified Pouchitis Disease Activity Index (mPDAI) score being less than 5 | Prospective |
| Ricart et al | 7 | 2 | 38 | 4.3 | 5 | A complete response was defined as cessation of fistula(s) drainage and total closure of all fistula(s), or cessation of diarrhea, incontinence, and abdominal pain. | Retrospective |
| Robbins et al | 92 | 50 | 32 | 5 | Not reported | we evaluated whether IBD serology changed over time with disease diagnosis and examined whether there were differences in serology between patients who developed denovo CD versus patients who did not develop pouch inflammation | Prospective |
| Shen et al | 17 | 12 | 36 | Not reported | 2 | Complete clinical response was defined as resolution of symptoms. Partial clinical response was defined as improvement in symptoms. | Prospective |
| Weaver et al | 56 | 24 | 44.1 | Not reported | 12 | Clinical response or remission was judged by the treating physician's assessment at 6 months. | Retrospective |

| | | | | | | | |
|---|---|---|---|---|---|---|---|
| Ferrante et al | 28 | 14 | 26 | Not reported | Not reported | Clinical response was defined as complete in case of cessation of diarrhea, blood loss, and abdominal pain, and as partial in case of marked clinical improvement | Retrospective |
| Gionchetti et al | 12 | 7 | 32.6 | Not reported | Not reported | Remission was defined as a combination of a clinical PDAI score of 1. QOL was assessed using Inflammatory Bowel less or equal 2 | Prospective |
| Gregory et al | 83 | 38 | 42.1 | 4.9 | 12 | clinical response at any time point was determined through chart review as defined in a prior study by our group as a decrease in the number of bowel movement, abdominal pain, or fistula drainage | Retrospective |
| Kelly et al | 42 | 11 | 32.6 | 63 | 12 | Complete response was defined as symptomatic and endoscopic resolution with modified Pouchitis Disease Activity Index (mPDAI) <5. | Retrospective |
| Khan et al | 12 | 3 | 41 | Not reported | Not reported | modified Pouchitis Disease Activity Index (mPDAI).mPDAI is the 18-point pouchitis disease activity index consisting of three principal component scores: symptom (range, 0–6 points), endoscopy, (range 0–6 points), and histology (range, 2–6 points). | Case series |
| Kjaer et al | 13 | 2 | 40.1 | Not reported | 3 | Primary outcome was reduction in clinical pouchitis disease activity index (PDAI) of more than 2 at any time. | RCT |

| | | | | | | | |
|---|---|---|---|---|---|---|---|
| Ollech et al | 24 | 14 | 35.6 | Not reported | 12.6 | Outcomes included a change in the endoscopic subscore of the Pouchitis Disease Activity Index (PDAI), change in the ulcerated surface area, clinical response, and the number of bowel movements per 24 h. | Retrospective |
| Segal et al (ileitis) | 29 | 17 | 53 | Not reported | 21 | Outcomes included the continued presence of PPI following biologic therapy, pouch failure defined by the need for an ileostomy | Retrospective |
| Segal et al | 34 | 19 | 25 | Not reported | 9.3 | The primary outcome was the development of IFX failure defined by early failure to IFX or secondary loss of response to IFX. | Retrospective |
| Singh et al | 19 | 10 | 26.7 | Not reported | 3 | The primary outcomes were to assess the improvement or reduction in the mPDAI symptom (range: 0–6) and endoscopy (range: 0–6) subscores after 3 months of vedolizumab therapy for CARP. | Retrospective |
| Verstockt et al | 33 | 13 | 34.4 | 4.8 | 3.5 | The primary endpoint, short-term clinically relevant remission was assessed at week 14, and defined as mPDAI<5 and reduction in the mPDAI score with2 points from baseline | Retrospective |
| Viazis et al | 7 | 3 | 37.1 | 6.2 | 36 | Complete clinical response was defined as cessation of diarrhea, urgency, incontinence, blood loss and abdominal pain. | Prospective |

| | | | | | | | |
|---|---|---|---|---|---|---|---|
| Viscido et al | 7 | 3 | 30 | 4 | 2.5 | Clinical response was classified as complete, partial, and no response. Fistulae closure was classified as complete, partial, and no closure. The pouchitis disease activity index (PDAI) and quality of life (QoL) were also used as outcome measures. | Prospective |
| Barreiro-de Acosta et al (1) | 33 | 18 | 45 | Not reported | 13 | Complete response was defined as cessation of diarrhea and urgency and partial response as marked clinical improvement but persisting symptoms | Retrospective |
| Barreiro-de Acosta et al (1) | 8 | | 42 | Not reported | 13 | The short-term and mid-term efficacy of ADA was evaluated | Retrospective |

Supplemental Table 3.

| | **Selection** | | | **Comparability** | **Outcome** | | | |
|---|---|---|---|---|---|---|---|---|
| Study | **Representativeness of sample (maximum: one star)** | **Sample size (maximum: one star)** | **Assessment of the exposure (maximum: one star)** | **Comparability of cohorts on the basis of the design or analysis (maximum: 2 stars)** | **Assessment of the outcome (maximum: one star)** | **Was follow up long enough? (maximum: one star)** | **Adequacy of follow up cohorts (maximum: one star)** | **Total score (maximum: 8 stars)** |
| Bar et al | * | * | * | ** | * | | | ****** (6) |
| Calabrese et al | * | | * | | * | | | *** (3) |
| Ferrante et al | * | * | * | * | * | * | | ****** (6) |
| Gregory et al | * | * | | ** | * | | | ***** (5) |
| Kelly et al | * | * | * | ** | * | | | ****** (6) |
| Ollech et al | * | * | * | | * | * | * | ****** (6) |
| Khan et al | * | | * | * | * | * | | ***** (5) |
| Segal et al | * | | * | ** | | * | * | ****** (6) |
| Segal et al (prepouch ileitis) | | | * | * | * | | | ***(3) |
| Singh et al | | | * | | * | | | ** (2) |
| Versockt et al | | | * | | * | | | ** (2) |

| | | | | | | | | |
|---|---|---|---|---|---|---|---|---|
| Viscido et al | * | * | | | * | | | *** (3) |
| Viazis et al | * | * | * | | * | | | *** (3) |
| Gionchetti et al | * | * | | * | * | | | **** (4) |
| Barreiro-de Acosta et al | * | * | * | * | | | | **** (4) |
| Colombel et al | * | | * | * | * | | * | ***** (5) |
| Haveran et al | * | | * | * | | | | ***(3) |
| Robbins et al | * | | * | | * | | | ***(3) |
| Ricart et al | * | * | * | | | | | ***(3) |
| Li et al | * | * | * | * | * | | * | ****** (6) |
| Shen et al | * | * | * | * | * | | | *****(5) |
| Weaver et al | * | | * | * | * | | * | ***** (5) |

Supplemental Table 4. Risk of Bias Assessment

| **Study** | **Risk of Bias** |
|---|---|
| Bar et al | Moderate |
| Calabrese et al | Moderate |
| Ferrante et al | Low |
| Gregory et al | Low |
| Kelly et al | Low |
| Khan et al | Moderate |
| Ollech et al | Low |
| Segal et al (prepouch ileitis) | Low |
| Segal et al | Low |
| Singh et al | Moderate |
| Versockt et al | Low |
| Viscido et al | Low |
| Viazis et al | Low |
| Gionchetti et al | Low |
| Barreiro-de Acosta et al | Low |
| Barreiro-de Acosta et al | Low |
| Colombel et al | Moderate |
| Haveran et al | Low |
| Robbins et al | Low |
| Ricart et al | Moderate |

| | |
|---|---|
| Li et al | Low |
| Shen et al | Low |
| Weaver et al | Low |
| Kjaer et al | Low |

**Sensitivity Analysis**

*Supplemental Figure. 1 Sensitivity analysis By Country*

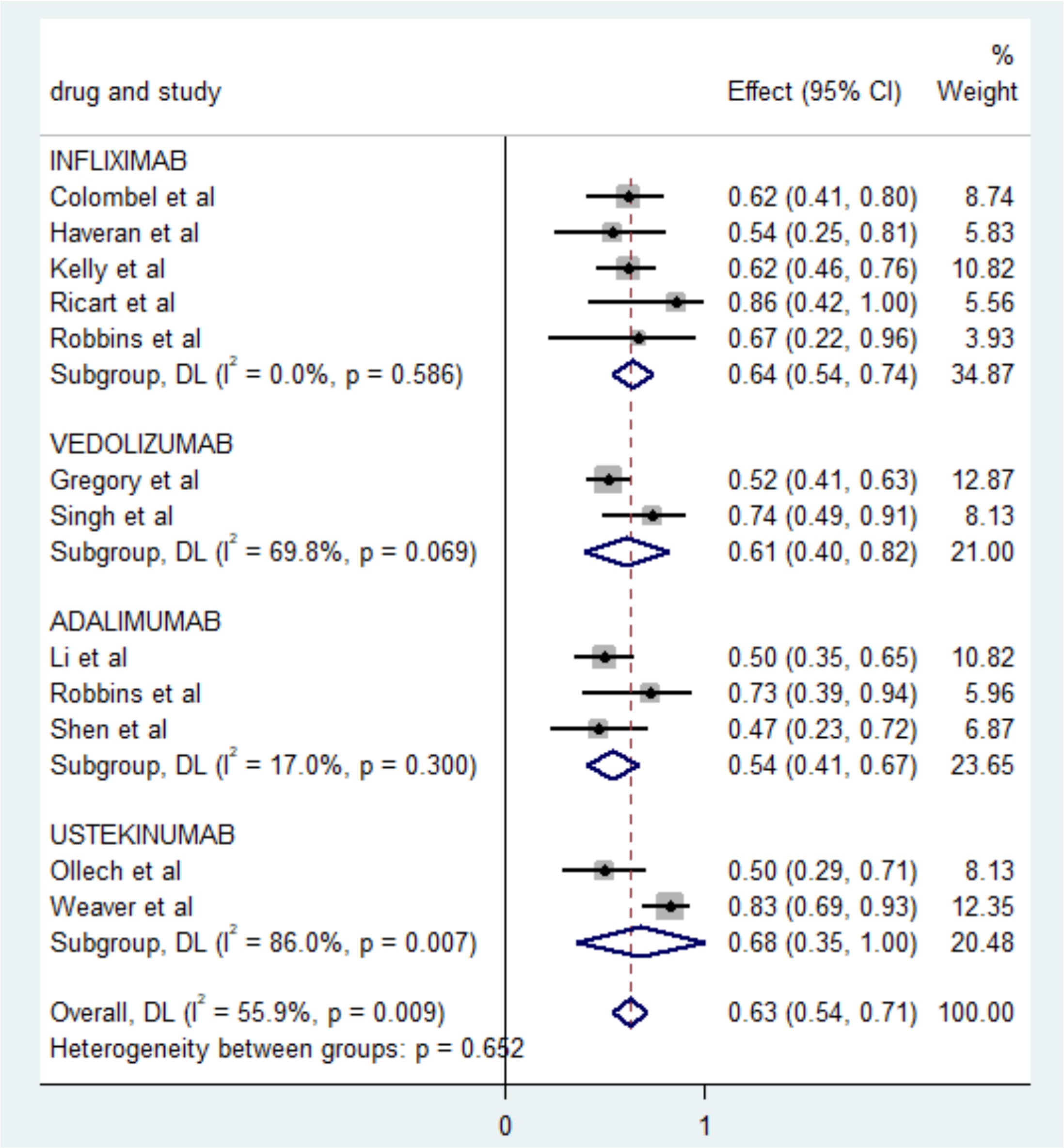

*Supplemental Figure. 2 Sensitivity analysis by quality of studies*

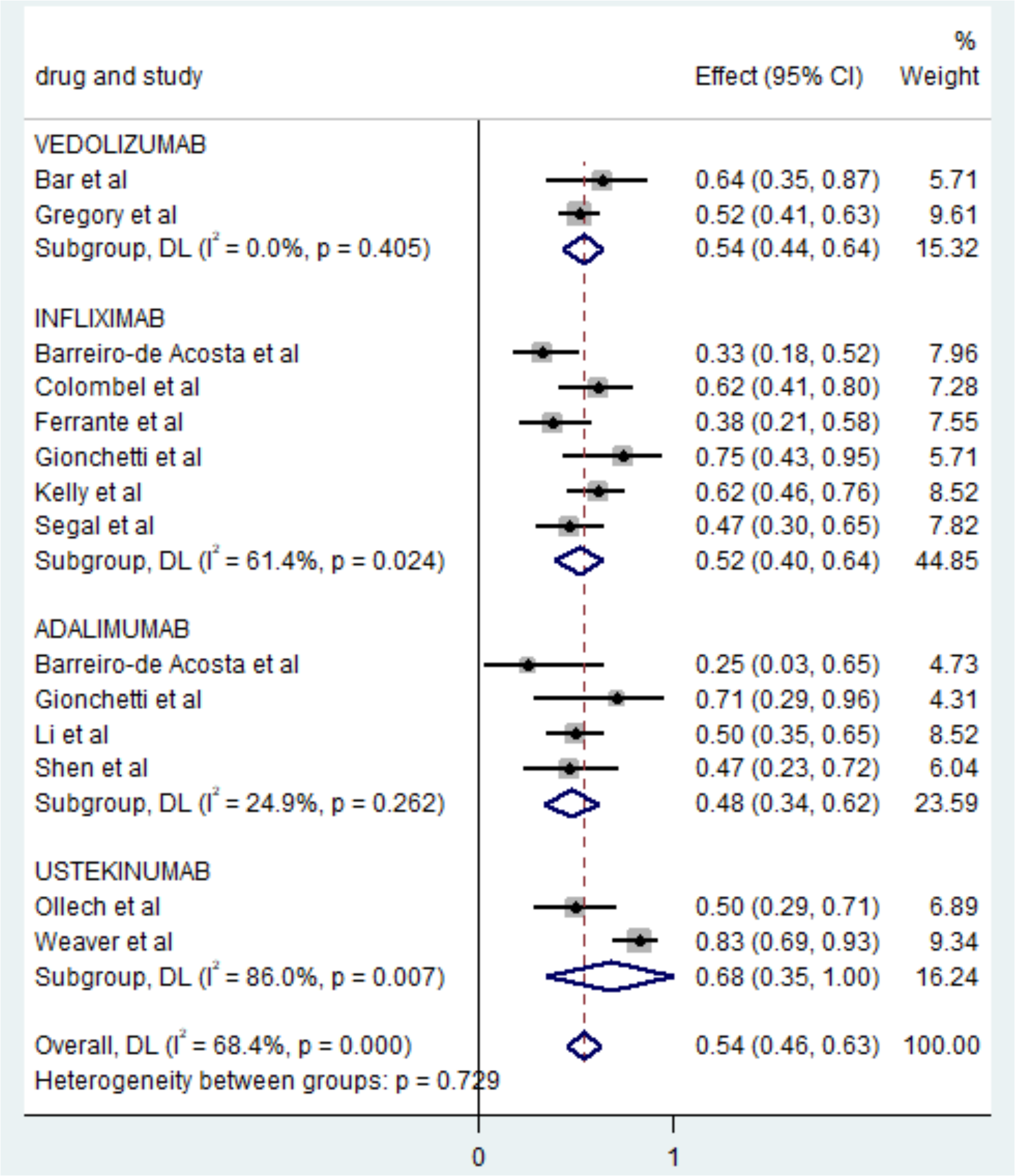

*Supplemental Figure. 3 Sensitivity analysis by year*

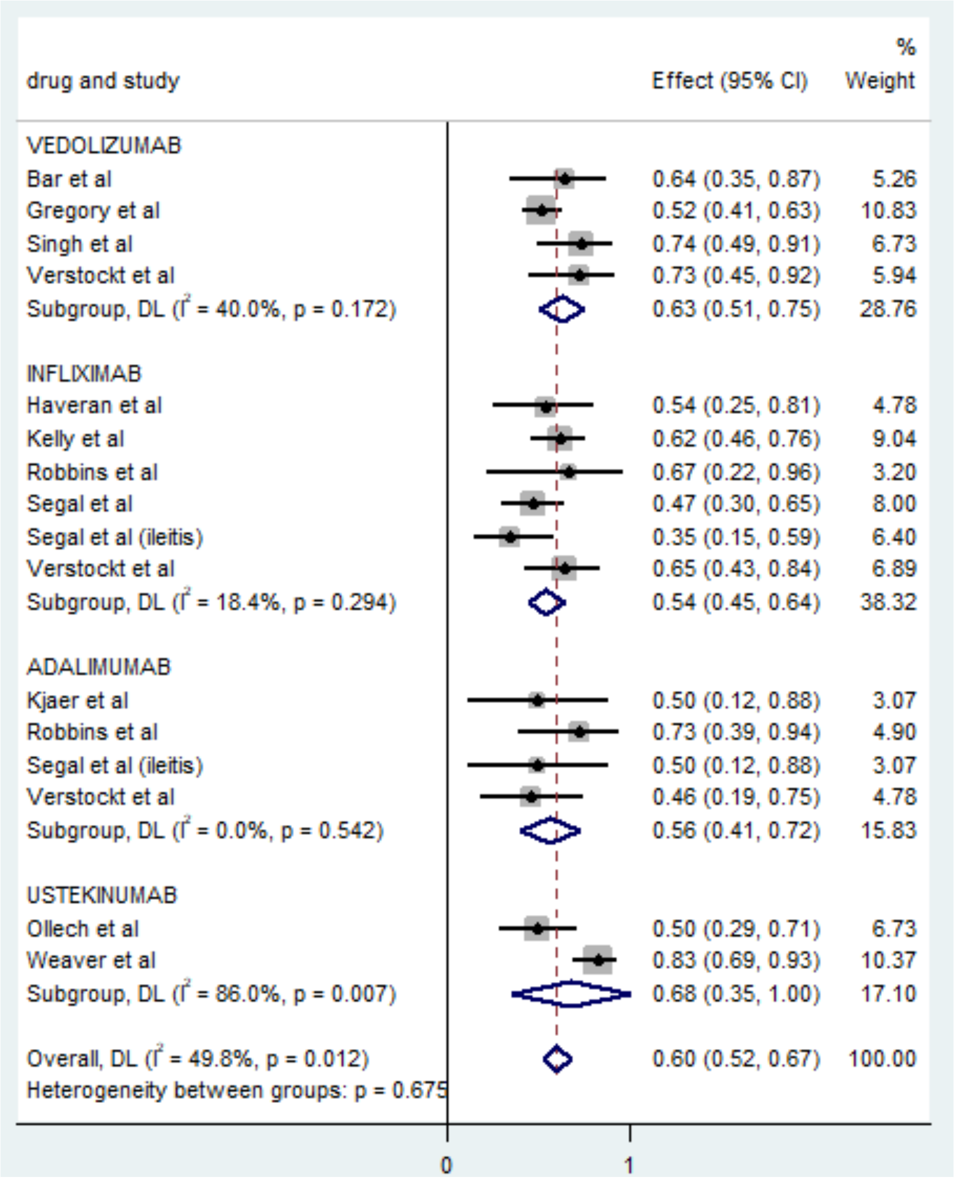

Supplemental Figure. 4 Publication Bias

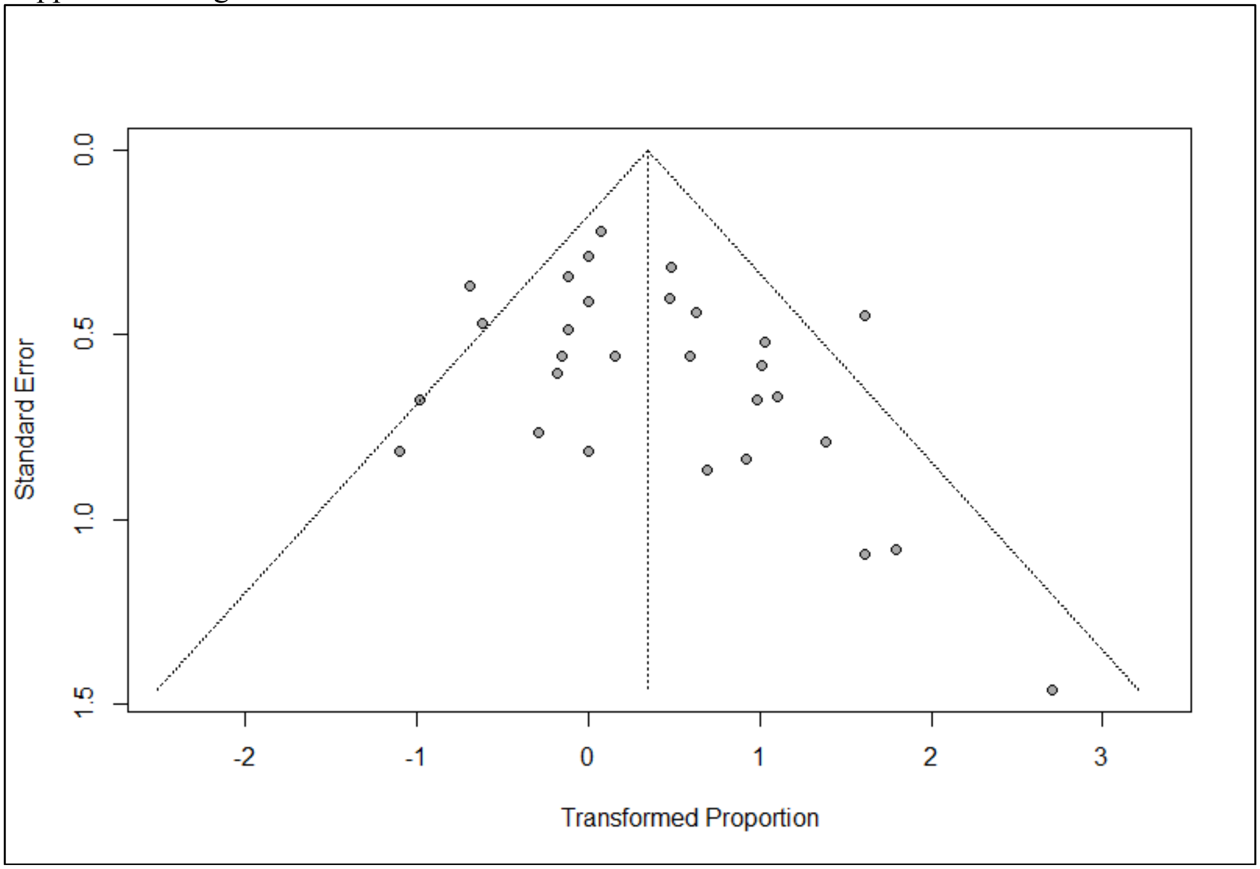
